# Bridging the Anesthesia Digital Data Gap in Low-Middle-Income Countries: Computer Vision-Ready Paper Health Records

**DOI:** 10.1101/2025.02.09.25321925

**Authors:** Bhiken Naik, Ryan Folks, Charbel Marche, Hannah Valenty, Matthew Beck, Lliam Brannigan, Christian Ndaribitse, Gregory Sund, Matthew Kynes, Bruce Biccard, Joshua Mann, Tracy Oyugi, Lisa Thannikary, Scott Jossart, Coreylyn Debettencourt, Donald E. Brown, Hyla Kluyts

**Author notes:** **<u>Corresponding Author:</u>** Bhiken I. Naik University of Virginia Health System Department of Anesthesiology **Mailing address: 1.** Hospital Dr Box # 4748, 1215 Lee Street, Charlottesville VA, 22903, USA.

## Abstract

**Introduction:** Surgical mortality is the third leading cause of death globally, with mortality rates in Africa double those of high-income countries despite patients being younger and undergoing lower-risk procedures. One of the contributors to poor outcomes in low- and middle-income countries (LMICs) is the lack of digital data, which is essential for quality improvement, audit and feedback systems, and early warning track-and-trigger systems. Due to limited financial resources, paper health records remain the standard in LMICs, making readily accessible digital data an urgent priority. This study builds on our previous work in computer vision-based digitization of smartphone-captured anesthesia records by developing a standardized, computer vision-ready anesthesia paper record. Designed for optimal digitization, this record will align with the Minimum Dataset for Surgical Patients in Africa guidelines.

**Methods:** The standardized, computer vision-ready anesthesia paper chart was developed with input from anesthesia experts in LMICs and data scientists. Key adaptations designed to facilitate accurate computer vision digitization included replacing traditional free-text entries with predefined categorical checkboxes, pre-printing the name of commonly used medications, supplementing handwritten medication names with numeric codes, and structuring input fields to improve computer vision digitization accuracy. Prior computer vision software was further iterated to improve digitization accuracy for the new standardized chart. Performance of the updated software running on the new computer vision-ready paper chart was then evaluated by comparing the software output to the human-annotated ground-truth data measuring both detection and interpretation accuracy.

**Results:** The training dataset consisted of thirty-three standardized, computer vision-ready anesthesia paper charts completed using synthetic data by a group of ten anesthesia providers. Five charts were reserved for validation, while the test dataset consisted of nine charts that were not used for any training or validation purposes. Updated computer vision software demonstrated high detection accuracy for vital signs: systolic blood pressure (93%), diastolic blood pressure (94%), and heart rate (93%), all physiological indicators (100%), and checkboxes (99%). The mean average error for inferring values from model detections were low: systolic (1.98mmHg), diastolic (1.13mmHg), heart rate (3.8/bpm), oxygen saturation (0.19%), end tidal carbon dioxide (0.65 mmHg), inspired oxygen concentration (2.48%). The accuracy for determining which checkboxes were marked vs. unmarked was 99%.

**Conclusion:** This study confirms the feasibility and accuracy of a standardized, computer vision-ready anesthesia chart that can be deployed in LMICs to facilitate digital data access.

## Introduction

Each year, an estimated 4.2 million people die within 30 days of surgery worldwide, with nearly half of these deaths occurring in low- and middle-income countries (LMICs).^1^ Surgical mortality is the third leading cause of death globally, following ischemic heart disease and stroke.^1^ According to the African Surgical Outcomes Studies, overall perioperative mortality in Africa is twice as high, pediatric perioperative mortality is 11 times as high, and maternal mortality is 50 times as high compared to high resource settings.^2, 3^

We believe a key contributor to poor surgical outcomes in LMICs is the lack of readily accessible digital data. Digital data plays a critical role in driving quality improvement and patient safety through audit and feedback systems, high-quality, context-specific research, and track-and-trigger systems designed to prevent ’failure to rescue’ events commonly observed in resource-constrained settings.^4–9^

One of the primary barriers to digital data availability in LMICs is the reliance on paper health records for documentation.^10^ Limited financial resources often prevent the widespread implementation of electronic health record systems and automated data collection tools, making paper records an indispensable part of healthcare delivery in LMICs.^11, 12^

Our recent efforts to create a comprehensive digital anesthesia dataset from two sub-Saharan African LMICs highlight the significant challenges encountered in converting analog paper records into digital data. Digitizing a single anesthesia chart required 15-30 minutes of manual annotation, depending on data complexity and volume. As an example, if a district hospital in sub-Saharan Africa performed 2,000-3,000 surgeries annually, this would translate to 50-75 weeks of full-time labor (assuming a 40-hour work week) to digitize all anesthesia records, a process that is both impractical and unsustainable.^13–15^

To bridge the digital data gap in LMICs, we previously demonstrated the feasibility of digitizing smartphone images of completed anesthesia paper health records using computer vision, a type of artificial intelligence.^16–22^ Our computer vision models achieved 99% accuracy for recognizing and classifying a checkbox that had been “checked” and a mean average error of 1.0 mmHg for systolic and 1.36 mmHg for diastolic blood pressure measurements when compared to human-annotated ground truth data.^17^

However, two major challenges remain. First, the accuracy for free-text features, such as medication type administration and numerical physiological parameters (e.g., pulse oximetry, tidal volume, blood loss, urine output), remains suboptimal at 36% and 85%, respectively, due to the difficulty of handwriting detection in computer vision, coupled with particularly illegible handwriting.^16, 17^ Second, the lack of a standardized anesthesia paper health record across hospitals poses a significant challenge. Each hospital uses its own unique anesthesia paper record, making the development and optimization of individual computer vision models for different chart formats resource-intensive and unsustainable.

Our goal is to design and develop a “standardized anesthesia paper health record”, which is computer vision ready for digitization. Target accuracy for this computer vision ready chart will exceed 90% for detecting marked checkboxes, symbol-denoted blood pressure and heart rate, medication data, and numerical physiological features when compared to ground-truth data. The design will incorporate essential data elements outlined in the ’Minimum Dataset for Surgical Patients in Africa’ guidelines, striking a balance between standardization and comprehensive clinical documentation tailored to the needs of LMICs.^23^

## Methods

### Study Approval

This study did not utilize any patient data to design, develop, or test the standardized computer vision-ready paper health record. All paper health records were populated exclusively with synthetic data. As a result, approval from both the Human and Social/Behavioral Sciences Institutional Review Board at the University of Virginia was waived.

### Data and Feature Engineering of Standardized Computer Vision Paper Adapted Anesthesia Charts

#### a. Data Elements

To ensure the standardized computer vision-ready anesthesia paper health record adhered to the “Minimum Dataset for Surgical Patients in Africa” guidelines, input on essential data elements was collected from anesthesia experts working in LMICs. The selection of data elements, guided by the standards developed by Kluyts et al., prioritized relevance and alignment with regional practices.^23^

#### b. Engineering of the Computer Vision Adapted Chart

Once consensus was obtained regarding the included data elements, the data science team designed and engineered the anesthesia paper health record to optimize digitization accuracy. Feature modifications and adaptations are demonstrated in Figure 1 and includes:

1. Converting as many free-text elements into categorical checkbox options (e.g., representing ASA Physical Status with six checkboxes instead of allowing free-text entries)
2. Pre-printing the names of commonly administered anesthesia medications used in LMICs.
3. Utilizing numeric codes paired with an accompanying data dictionary to enable the identification of medications not included in the pre-printed list.
4. Where possible, a numeric entry that is segmented and enclosed within its own designated box to enhance digit recognition accuracy.
5. Incorporating clearly defined numeric timestamp fields to ensure precise data capture for surgery and anesthesia durations.

**Figure 1:**
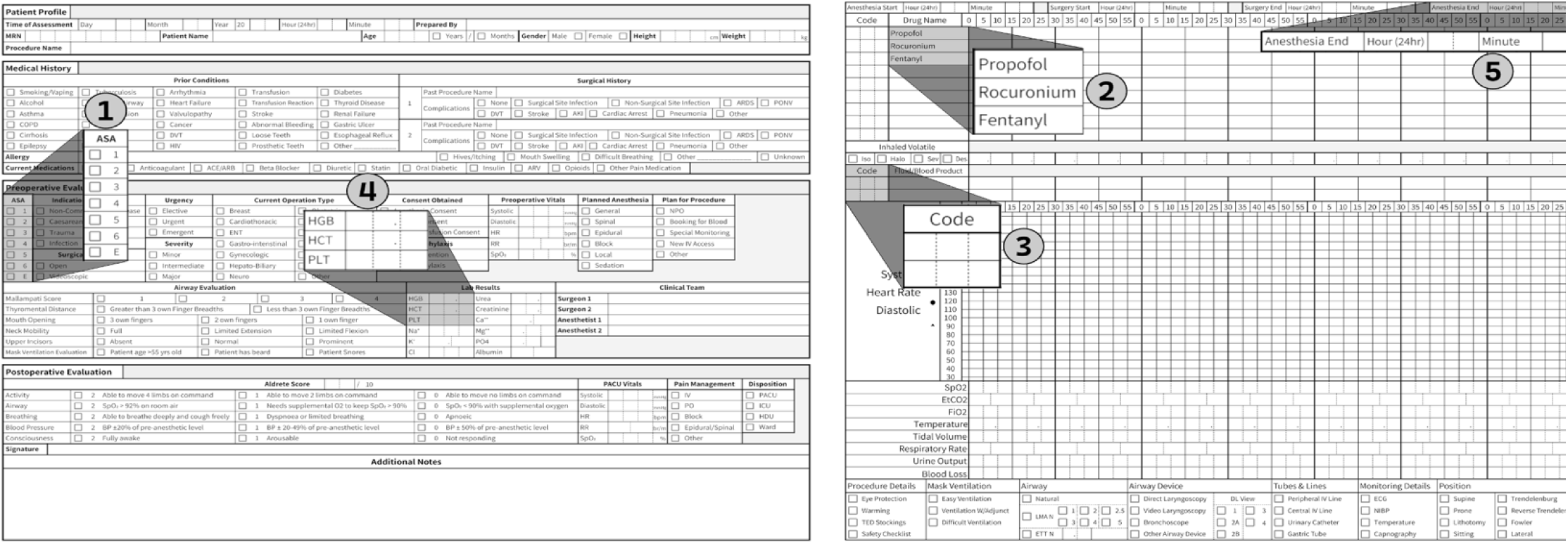
Feature Modifications in Computer Vision Ready Anesthesia Paper Record. 1-Coverting free text to checkboxes, 2-Pre-printed medication, 3-Numeric code to identify medications and fluids, 4-Segmented numeric entry, 5-Numeric timestamps

### Creation of Synthetic Computer Vision Adapted Anesthesia Charts

After finalizing the standardized computer vision-ready anesthesia chart, a diverse group of ten US-based anesthesia providers, including both trainees and attending anesthesiologists, were tasked with completing 4-5 of these charts each. This sample size was determined by balancing the necessity to capture enough data in diverse handwriting styles with the time expenditure of manually annotating all the bounding box data for a single chart. Providers were instructed to use fictitious, synthetic data while varying pen colors and handwriting styles to introduce heterogeneity in text features. This approach ensured the generation of a robust and diverse dataset for evaluating the chart and model performance in real-world scenarios.

### Model Iteration

#### Prior Work

Our prior work established a framework for digitizing anesthesia records using smartphone images and computer vision analysis.^16–19, 22^ Preprocessing techniques are applied to enhance image quality and align charts for improved data extraction. YOLOv8 object detection models identify and classify key physiological parameters, including blood pressure, heart rate, and checkboxes. Clustering algorithms further refine digit recognition, ensuring accurate data grouping and classification. The data extracted by our software from the smartphone images aligned closely with ground-truth data, with low mean average errors for blood pressure and heart rate measurements. Additionally, checkbox classification achieves high precision, effectively distinguishing marked from unmarked selections.

#### Model Updates for the Standardized Computer Vision-Ready Chart

We updated our previous computer vision software to align with the new standardized computer vision-ready paper chart and improve accuracy. Specific computer vision software developments included:

##### 1. Pose Estimation for Symbol-denoted Blood Pressure

Previously, our models estimated blood pressure by determining the y-coordinate of the top of the bounding box and calculating a second y-coordinate based on a fixed proportion of the box’s height (90% for systolic, 10% for diastolic), then finding the point on the chart’s blood pressure legend that was closest. Now, by leveraging a Pose detection model, a bounding box that encompasses the entire blood pressure symbol as seen in Figure 2, and the exact point that marks the ‘tip’ of the blood pressure arrow can be detected simultaneously, yielding higher precision.^24^ Similarly, we utilized the Pose model for heart rate data.

**Figure 2:**
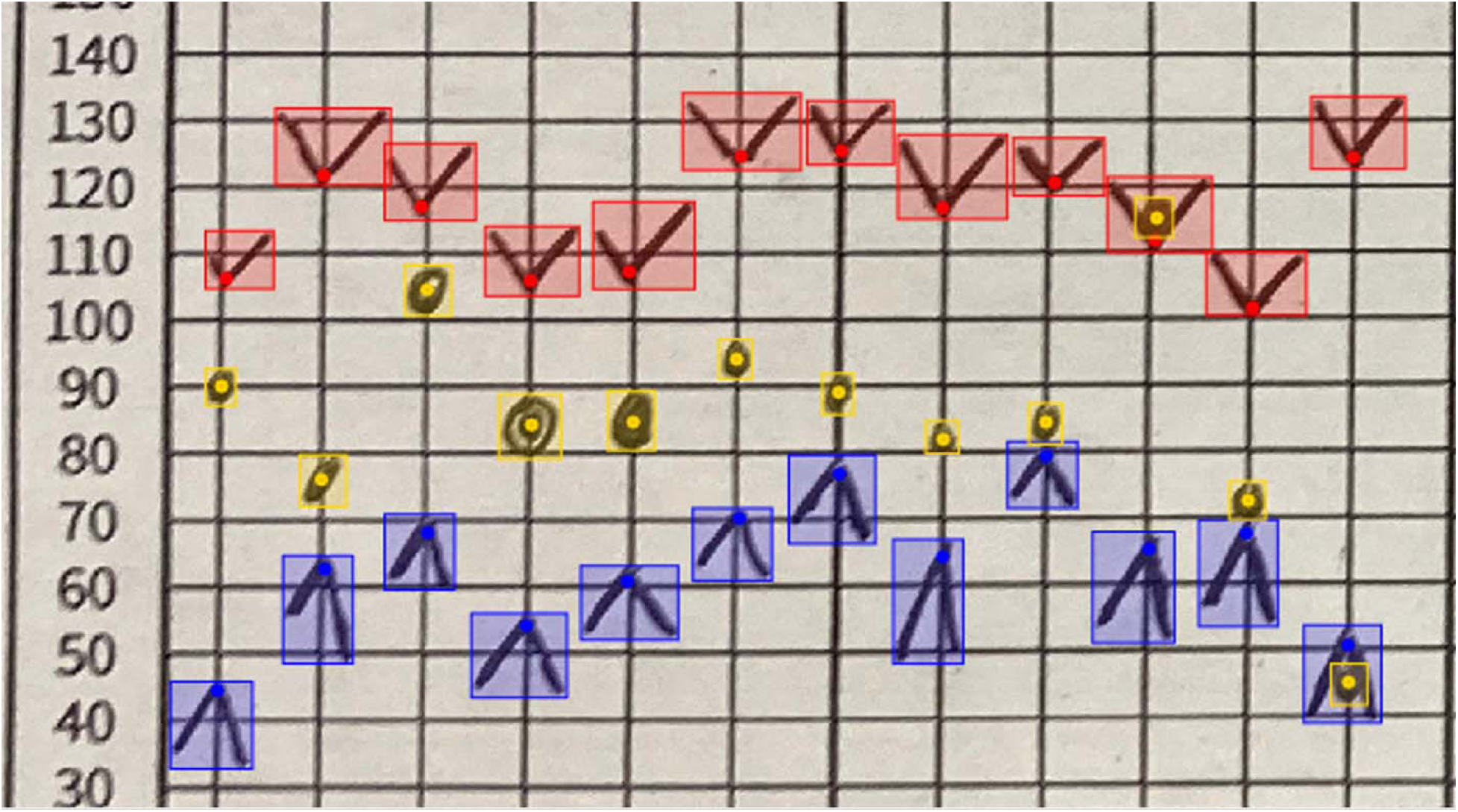
Bounding box and keypoint annotations on blood pressure and heart rate symbols. Bounding boxes are rectangle annotations that are just large enough to include the entire object within them. Keypoints are points inside bounding boxes that correspond to areas of interest on the object.

##### 2. Exact Timestamp Estimation for Blood Pressure and Physiological Indicators

Previously, the model sorted the time detections from left to right and assumed the leftmost detection to be at the 0-minute mark. The updated model detects all the printed digits that form the timestamp legend that runs left to right along the top of the blood pressure section, then clusters those digits together to create exact timestamp locations and uses this to find the timestamp for each blood pressure and heart rate symbol. Using this timestamp legend, timestamps for physiological indicators can also be determined.

##### 3. Predefined Boxes for Handwritten Digits

The standardized computer vision-ready chart requires anesthesia providers to write digits within predefined boxes, as demonstrated in Figure 1, point 4. This structured format improves digitization accuracy for physiological indicators and allows for many more data elements to be digitized, such as surgical timing, laboratory values, and demographic data.

### Model Performance Analysis

Prior to performing accuracy analysis of the updated standardized computer vision-ready paper chart and associated models, we created a human-annotated ground-truth dataset for all the synthetic charts created. Data was manually extracted by a single reviewer and recorded in an excel spreadsheet for comparison to model outputs.

### Accuracy of Computer Vision Models

To determine the accuracy of our computer vision models, we reported metrics in two stages, detection accuracy and interpretation accuracy. Detection refers to the ability of the model to find the objects of interest in the standardized computer vision-ready paper chart. Interpretation refers to the ability of the model to create a meaningful interpretation of the objects that have been detected.

#### Detection Accuracy

Detection accuracy metrics were reported as accuracy, precision, recall, and F1. They were computed as outlined, where TP, FP, FN, TN are the number of true positives, false positives, false negatives, and true negatives, respectively.

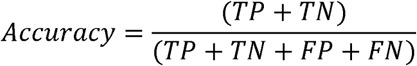

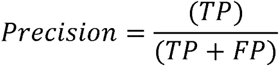

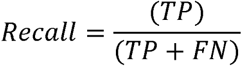

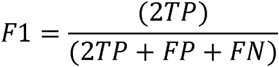

Detection accuracy was reported for systolic, diastolic blood pressure and heart rate symbol data. For checkboxes, both detection and correct interpretation of whether a checkbox was marked were reported using the detection accuracy metrics.

#### Interpretation Accuracy

For interpretation accuracy metrics, mean average error was used for blood pressure, heart rate, and physiological data. Mean average error (MAE) is computed as the average of the differences between the model extracted and the ground-truth values.

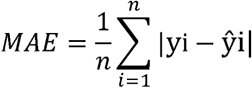

where n is the number of items, y_i_ is the ground-truth value and ŷ_i_ is the model extracted values. All software developed was written in the Python3 programming language and are freely available under the GPL 3.0 license. The computer vision models were trained using the Ultralytics package, and all software developed for this study is publicly available on our GitHub repository for reproducibility and further validation.^25–27^

## Results

### Training & Testing Datasets

The training dataset consisted of thirty-three standardized computer vision-ready paper charts, populated with synthetic data by a group of ten clinicians, to obtain a variety of handwriting qualities and styles. The data-points generated were all within plausible biological ranges and varied in case-length, resulting in some charts having more data-points than others. Five standardized computer vision-ready paper charts were reserved for validation. The test dataset consisted of nine standardized computer vision-ready paper charts that were not used for any training or validation purposes. All detection and interpretation accuracy are reported for the test dataset only.

### Detection Accuracy

Detection of the systolic, diastolic, and heart rate symbols within the standardized computer vision-ready paper charts are reported in Table 1, while checkbox detection metrics are reported in Table 2. Detection accuracy was consistently above 93% for all relevant data elements.

**Table 1:**
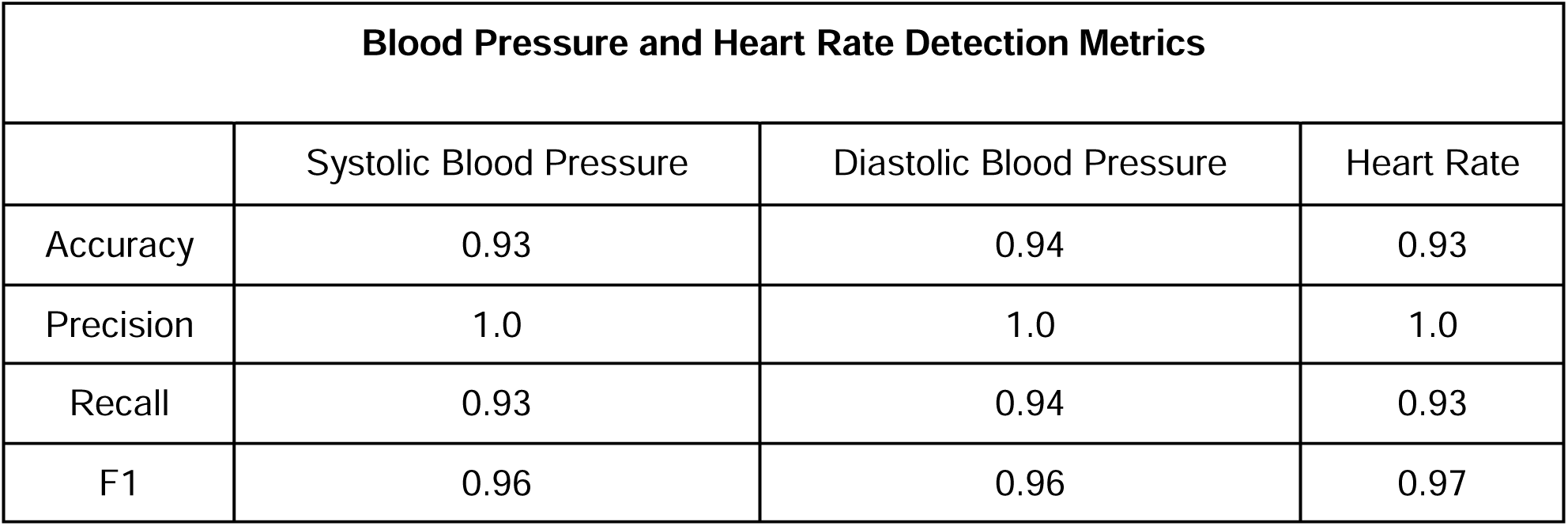
Detection metrics for blood pressure and heart rate. These metrics report whether a blood pressure or heart rate symbol was correctly identified for a given timestamp.

| Blood Pressure and Heart Rate Detection Metrics |  |  |  |
| --- | --- | --- | --- |
|  | Systolic Blood Pressure | Diastolic Blood Pressure | Heart Rate |
| Accuracy | 0.93 | 0.94 | 0.93 |
| Precision | 1.0 | 1.0 | 1.0 |
| Recall | 0.93 | 0.94 | 0.93 |
| F1 | 0.96 | 0.96 | 0.97 |

**Table 2:** Detection metrics for checkboxes, refer to whether a checkbox was detected in the correct location.

| Checkbox Detection Metrics |  |
| --- | --- |
| Accuracy | 0.99 |
| Precision | 1 |
| Recall | 0.99 |
| F1 | 0.99 |

### Interpretation Accuracy

Interpretation accuracy for blood pressure, heart rate, and numerical physiological data are reported in Table 3. The ability of the model to accurately determine whether a checkbox is marked or not is reported in Table 4. Finally, laboratory data detection and interpretation metrics are reported in Table 5. The mean average error was extremely small for all variables of interest, while the model’s ability to identify whether a checkbox was marked or not was consistently 99%.

**Table 3:** Interpretation accuracy for data variables reported as mean average error compared to the human-annotated, ground-truth dataset. Sp02-Oxygen saturation, EtC02: End-tidal carbon dioxide, Fi02-Inspired oxygen concentration.

| **Data Variable** | **Mean Average Error** |
| --- | --- |
| Systolic Blood Pressure | 1.98 mmHg |
| Diastolic Blood Pressure | 1.13 mmHg |
| Heart Rate | 3.78 bpm |
| SpO2 | 0.19 % |
| EtCO2 | 0.65 mmHg |
| FiO2 | 2.48 % |
| Temperature | 0.32 °C |
| Tidal Volume | 11.66 ml |
| Respiratory Rate | 0 br/m |
| Urine Output | 0 ml |
| Blood Loss | 3.5 ml |

**Table 4:** Interpretation metrics for checkboxes. Metrics describe model accuracy to correctly classify a checkbox detection as checked vs. unchecked.

| Checkbox Interpretation Metrics |  |
| --- | --- |
| Accuracy | 0.99 |
| Precision | 0.99 |
| Recall | 0.99 |
| F1 | 0.99 |

**Table 5:** Laboratory value interpretation metrics. Interpretation metrics are reported as correct divided by total number of laboratory tests in the dataset, where correct occurs when the predicted lab value is exactly equal to the true lab value. Hgb-Hemoglobin, Hct-Hematocrit, Plt-Platelets, Na-Sodium, Cl-Chloride, Ca-Calcium, Mg-Magnesium, P04-Phosphate.

| Laboratory Value Interpretation Metrics |  |  |  |
| --- | --- | --- | --- |
| Lab Value | Correct | Total # in Test Set | Percent Correct |
| Hgb | 5 | 6 | 0.83 |
| Hct | 5 | 6 | 0.83 |
| Plt | 5 | 6 | 0.83 |
| Na | 6 | 6 | 1.0 |
| Cl | 6 | 6 | 1.0 |
| Urea | 6 | 6 | 1.0 |
| Creatinine | 6 | 6 | 1.0 |
| Ca | 5 | 5 | 1.0 |
| Mg | 5 | 5 | 1.0 |
| P04 | 4 | 5 | 0.8 |

## Discussion

Our work demonstrates the feasibility of a standardized computer vision-ready paper chart that enhances the digitization accuracy of computer vision models while aligning with the ’Minimum Dataset for Surgical Patients in Africa’ guidelines. This work represents an innovative, low-cost, and scalable solution to improve the anesthesia digital data gap in LMICs, which is unlikely to improve soon, due to the lack of financial resources to implement and maintain electronic health records. Access to high-quality digital data can catalyze quality improvement and research in LMICs, which are critical to enhancing patient outcomes.

As an example, while anesthesia-related mortality in HICs has declined from 684 per million in the 1970s to 85 per million by the 2000s, mortality rates in LMICs have increased from 326 per million to 467 per million over the same period.^2^ The increased mortality in LMICs attributed to anesthesia care represents a convergence of three key issues: insufficient number of trained anesthesia providers, inadequate equipment for monitoring during surgery, and suboptimal quality of anesthesia care, characterized by a critical failure to follow evidence-based best practice standards.^3,30–32^

Quality of anesthesia care can be improved in LMICs by implementing audit and feedback systems, which have been shown to improve adherence to standards of care and reduce postoperative complications. However, audit and feedback are predicated on access to high-quality digital data, which is problematic in hospitals that use anesthesia paper health records. Implementing the standardized computer vision adapted anesthesia paper health record with the digitization model developed by our team affords a novel approach for anesthesia providers in LMICs to have site-specific quality data that can be leveraged to identify gaps in anesthesia care and implement quality improvement measures.

The absence of digital data also significantly hinders the ability to conduct high-quality research in LMICs. Currently, most anesthesia research originates from HICs, with findings often inappropriately extrapolated to LMICs. This overlooks critical nuances such as demographic, socioeconomic, and geopolitical factors that uniquely affect surgical outcomes in these regions. Digitizing standardized anesthesia paper health records can address these challenges by harmonizing research data elements across institutions in LMICs, enabling high-quality, multicenter observational research tailored to their unique needs.

To ensure the scalability of our approach, future work will focus on developing a mobile health (mHealth) platform that integrates computer vision models for point-of-care use by anesthesia providers. This platform will streamline key tasks, including capturing a smartphone image of the completed anesthesia paper health record, uploading it to the app, digitizing the anesthesia paper health record, and securely storing the data in a cloud-based relational database, all within a seamless workflow.

Finally, the computer vision models and standardized anesthesia paper health record dyad developed in this study has potential applications beyond anesthesia care. Intensive care unit paper health records share many similarities with anesthesia charts, including physiological data elements, symbol-denoted blood pressure and heart rate data, medication administration records, categorical data fields, and free-text sections. By extending our approach to critical care settings, this system could generate robust digital datasets that support early warning and track-and-trigger systems, such as the Modified Early Warning Score (MEWS), enhancing patient monitoring and clinical decision-making in critical care units in LMICs.

This study has several limitations, which will need to be addressed to ensure broad acceptability by local stakeholders. First, anesthesia experts from three sub-Saharan countries, South Africa, Kenya, and Rwanda, were recruited to develop the standardized computer vision adapted anesthesia paper health record. The chart developed may not be generalizable to other countries, who may have institutional, regional, or national requirements for documentation. To mitigate this risk we used the “Minimum Dataset for Surgical Patients in Africa” guidelines to develop the chart and ensure key data elements suggested by this guideline were included. In addition, the current standardized chart is written only in English. With a multitude of languages spoken across LMICs, these charts will need linguistic alignment for broader acceptability. However, this should be a relatively straightforward problem to solve, as symbol-denoted features are universally utilized, data dictionaries with associated numerical codes can be modified, and categories associated with checkboxes can be updated to the language preference of choice for the anesthesia provider using a reverse translation approach. Finally, while the utilization of checkboxes and standardized data entry simplifies data collection, it cannot capture the subtlety and complexity of a comprehensive patient evaluation, anesthetic plan, and perianesthetic course. This limitation is inherent in any health record, and the trade-off between data granularity and ease of use may require some institutions to supplement this record with additional systems to document patient care.

## Conclusion

This study addresses the critical gap in digital anesthesia data in LMICs, where reliance on paper health records hinders quality improvement, research, and patient safety initiatives. By designing a standardized anesthesia paper health record adapted for computer vision digitization, we demonstrated excellent accuracy while adhering to the “Minimum Dataset for Surgical Patients in Africa” guidelines. The proposed solution is scalable, low-cost, and adaptable, offering a pathway to generate robust digital datasets for quality improvement, research, and clinical decision-making in resource-limited settings. Future work will focus on expanding this approach to other clinical areas and integrating it into an mHealth platform for point-of-care use.

## Supplementary Content

Supplementary Table 1: Descriptive statistics of synthetic data recorded in the computer vision-ready paper health record for the test set. All data reported as the number of specific data elements in the test set and median[interquartile range] or % for that variable.

**Clinical Trial Number / Registry URL:** Not applicable

**Prior Presentations:** This work has not been presented previously

**Funding Statement:** This work is funded in part from a grant from the Center for Global Inquiry and Innovation, University of Virginia.

**Conflict of Interest:** No conflicts of interest reported by any authors

## Data Availability

All software developed for this study is publicly available on our GitHub repository for reproducibility and further validation

## Notes

### Competing Interest Statement

The authors have declared no competing interest.

### Clinical Protocols

https://github.com/Paper-Chart-Extraction-Project/ChartExtractor

