## Supplementary Table 1 for "Bridging the Anesthesia Digital Data Gap in Low-Middle-Income Countries: Computer Vision-Ready Paper Health Records"

| **Test Set Descriptive Statistics** | |
| --- | --- |
| **Variable Name (Number of Data)** |  |
| Systolic Blood Pressure (mmHg) (n=135) | 127 [15, 147] |
| Diastolic Blood Pressure (mmHg) (n=135) | 73 [62, 86] |
| Heart Rate (bpm) (n=132) | 101 [89, 123] |
| Oxygen Saturation (%) (n=61) | 99 [98, 100] |
| End Tidal Carbon Dioxide (mmHg) (n=51) | 37 [34, 39] |
| Fraction of Inspired Oxygen (%) (n=52) | 40 [32, 65] |
| Temperature (°C) (n=33) | 36.2 [35.9, 36.9] |
| Respiratory Rate (/min) (n=39) | 14 [12, 16] |
| Urine Output (ml) (n=20) | 78 [38, 121] |
| Blood Loss (ml) (n=17) | 300 [200, 600] |
| Checkboxes N (%) | Checked=121 (29), Unchecked=302 (71) |
| Hgb (g/L) (n=6) | 12.3 [10, 15] |
| Hematocrit (%) (n=6) | 36 [26, 51] |
| Platelets (^9/L) (n=6) | 237 [138, 324] |
| Na (mEq/L) (n=6) | 140 [138, 148] |
| K (mEq/L) (n=6) | 4.1 [3.4, 4.6] |
| Cl (mEq/L) (n=6) | 104 [100, 110] |
| Urea (mg/dl) (n=6) | 8.4 [5.5, 12.7] |
| Creatinine (n=6) | 12.5 [3.4, 19.4] |
| Ca  (mg/dl) (n=5) | 2.4 [2.3, 5.7] |
| P04 (mg/dl) (n=5) | 1.2 [1.2, 2.1] |
| Albumin (g/dl) (n=3) | 27 [25, 30] |

Supplementary Table 1: Descriptive statistics of synthetic data recorded in the computer vision-ready paper health record for the test set. All data reported as the number of specific data elements in the test set and  median[interquartile range] or % for that variable .
